# A Precision Aging Approach to Cognitive Aging: Clustering Multiple Domains of Risk

**DOI:** 10.64898/2026.08.18.26360626

**Authors:** Lee Ryan, Olivia S. Ortiz, Corinne A. Pettigrew, Anya Soldan, Bonnie LaFleur, Bonnie Levin, Tatiana Rundek, James J. Lah, Meredith Hay, Abhay Moghekar, Kristian P. Doyle, Carol A. Barnes, Matt J. Huentelman

**Affiliations:** Departments of Psychology, Neurology, Neuroscience and Radiology and Imaging Science,,University of Arizona, Tucson, AZ, USA; Department of Neurology, Johns Hopkins University, Baltimore, MD, USA; Department of Neurology, University of Miami, Miami, FL, USA; Department of Neurology, Emory University, Atlanta, GA, USA; Translational Genomics Research Institute, Early Detection and Prevention Division, Phoenix, AZ, USA

**Keywords:** precision aging, risk factors for cognitive and health outcomes, cluster analysis, memory performance, profiles of risk

## Abstract

The term *Precision Aging* describes an approach that focuses on multi-domain profiles of risks impacting individual trajectories of age-related cognitive functioning. The goal of the present study was to identify profiles of risk within a sample of 555 adults, ages 50 to 79, without diagnosis of dementia. Using cluster analyses, we considered 38 risk factors associated with five categories of risk known to negatively impact cognitive aging – cardiovascular insufficiency, glucose dysregulation, inflammation, immune dysfunction, and neuropathology. Results yielded five profiles, including a group with low risk in all five risk categories, and four groups with prominent risks in specific domains. Importantly, all four high risk groups performed more poorly relative to the low risk group on multiple memory measures from a well-established neuropsychological test, the Auditory Verbal Learning Test. The results highlight the importance of considering multiple domains of risk within the same cohort to predict age-related cognitive functioning.

## Introduction

Numerous factors are associated with age-related cognitive impairment, cognitive decline, and increased risk for dementia, including unhealthy behaviors such as physical inactivity and poor diet, chronic conditions including heart disease, hypertension, and diabetes, and psychosocial stressors including social isolation, among many others. But each risk factor, in isolation, applies to only a subset of individuals and explains only a small portion of variance in cognitive functioning. Ryan et al. (2019) applied the term *Precision Aging* to describe an individualized approach that focuses on multi-domain profiles of risks and resilience impacting cognitive functioning. Two older adults experiencing cognitive impairment may have very different risks – one with cardiovascular dysfunction due to unhealthy lifestyle, the other with systemic inflammation due to chronic stress. The most effective treatment for these two individuals will likely be quite different, based not on their cognitive functioning, but on their profile of risks. A precision aging approach is critical for developing precise and effective individualized interventions to maintain brain and cognitive health across the lifespan.

Cluster analysis is one method that has been used to identify age-related profiles of risk and their differential impact on cognitive and health outcomes. Most commonly, clustering algorithms have been applied to multiple measures within a single domain of risk, such as frailty (Chew al., 2021), sleep metrics (Djongalic et al., 2021), and lifestyle factors (Roca-Ventura et al., 2025). Few studies have considered multiple domains of risk within the same cohort of older adults. In one study, Sebastiani et al. (2017) clustered standard blood measures, lipid and inflammatory biomarkers, and measures of physical frailty. Several patterns were associated with higher risk of future cancer, cardiovascular disease, type-2 diabetes, and all-cause mortality. Although cognition was not assessed, a critically important finding was that individual biomarkers had limited predictive value, highlighting the importance of considering multiple biomarkers within and across categories of risk.

In the present study, we consider five major risk categories associated with age-related cognitive impairment, reviewed in Ryan et al. (2019). ***Cardiovascular insufficiency*** includes highly prevalent and often co-morbid conditions, such as obesity, hypertension, and heart failure which have an additive effect on risk for cognitive impairment and cognitive decline (Lewis et al., 2021; Kaffashian et al., 2011). ***Glucose dysregulation*** includes diabetes and prediabetes, a condition characterized by elevated fasting glucose levels and elevated HbA1c levels in the absence of diabetes. Among middle-aged and older adults, glucose dysregulation is a risk factor for cognitive impairment and dementia (Rawlings et al., 2014; Roberts et al., 2014). Increased ***inflammation*** occurs in the aging brain, with peripheral markers such as interleukin-1alpha (IL-1α) and interleukin 6 (IL-6) linked to decreased cognitive performance (Teunissen et al., 2003; Leonardo et al., 2023). Closely related, increased production of proinflammatory cytokines reflecting age-related ***immune dysfunction*** is associated with poorer cognition among older adults (Farina et al., 2022), even after adjusting for other factors such as *APOE* status and cardiovascular risk (Fang et al., 2022). Finally, there is growing interest in biomarkers of ***neuropathology*** and their association with age-related cognitive functioning. In individuals without dementia, Aβ plaque accumulation and phosphorylated tau are associated with poorer cognition cross-sectionally and increased rate of cognitive and clinical decline (Soldan et al., 2025; Sperling et al., 2019). Other biomarkers, including neurofilament light chain protein (NfL) reflecting neuronal damage (Palmer et al., 2023), and glial fibrillary acidic protein (GFAP), reflecting astrocyte activation, also predict steeper cognitive decline among older adults (Saunders et al., 2023).

The goal of the present study was to identify profiles of risk for age-related cognitive impairment within a sample of 555 community-dwelling adults, ages 50 to 79, without a diagnosis of dementia, from the Healthy Minds for Life (HML) study in the Precision Aging® Network (PAN; https://precisionagingnetwork.org/). Using a two-step process of hierarchical clustering followed by k-means cluster analysis, we considered 38 individual risk factors associated with the five categories of risk described above – cardiovascular insufficiency, glucose dysregulation, inflammation, immune dysfunction, and neuropathology – and how the resulting profiles related to multiple measures of memory performance from a well-established neuropsychological memory test, the Auditory Verbal Learning Test.

## Results

### Risk category scores

Frequency distributions for each of the five risk categories shown in **Figure 1**. Most of the risk category scores were highly skewed, with approximately half the sample having no risk in a given category. The exception was cardiovascular health risks, which were relatively normally distributed with a mean of 2.83 risks out of a possible 10 (see **Figure 1**). Only 2.5% of participants had no cardiovascular risk, and another 8.6% of the sample had only 1 cardiovascular risk factor. The majority of the sample (71%) fell within the range of 2-4 cardiovascular risks, while 18.5% had five or more risks in this category.

**Figure 1.**
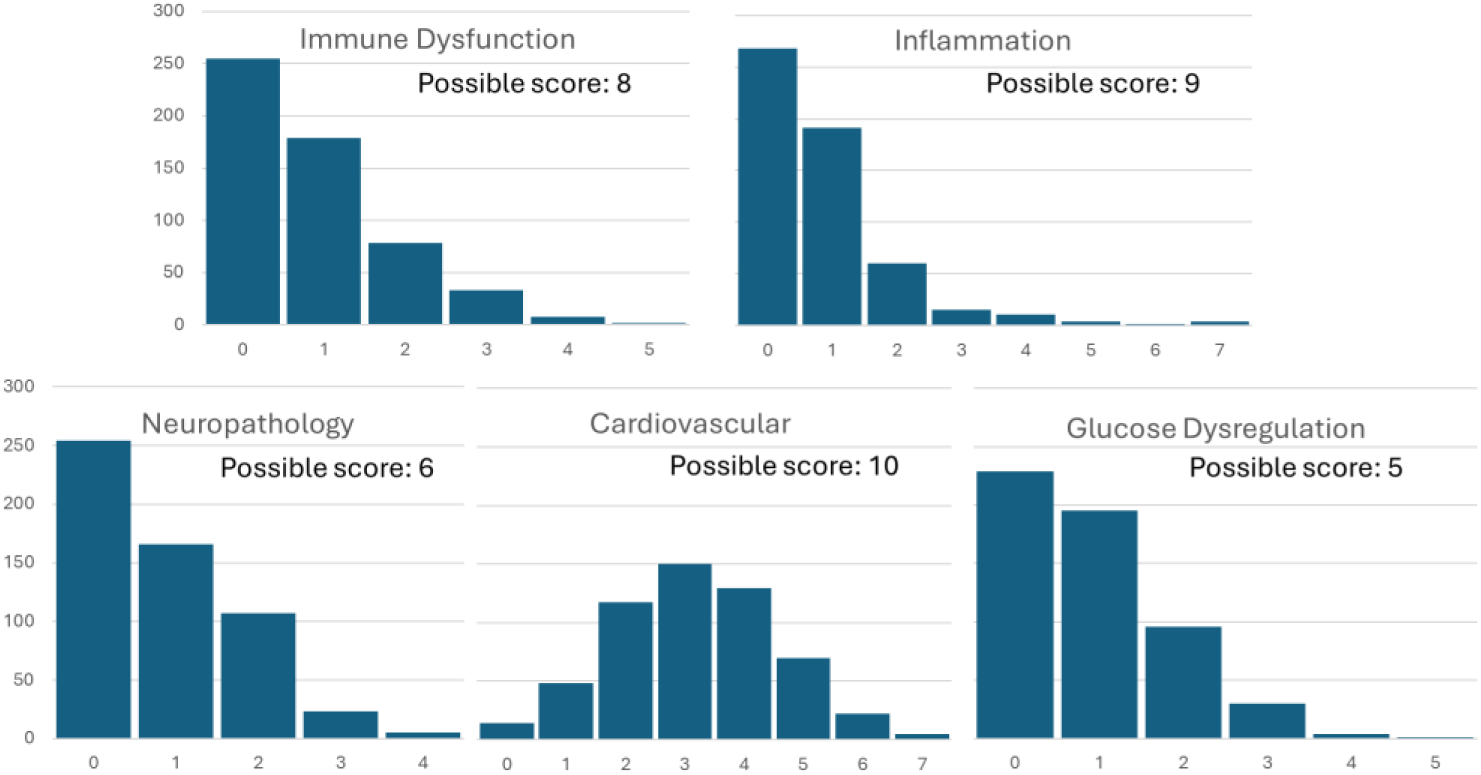
Figure 1 shows the frequency distribution of non-standardized scores for each of the five risk categories. ‘Possible score’ indicates the maximum score possible given the number of risk factors considered in the risk category.

### Determining the optimal number of clusters

The five standardized risk scores were entered into an agglomerative hierarchical cluster analysis using Ward’s linkage method and squared Euclidean distances. Visual inspection of the resulting dendrogram **(Figure 2, left panel**) suggested two possibilities – either a four cluster solution (highlighted in pink, green, blue, and rust boxes) or five cluster solution (separating the pink cluster into two separate smaller clusters, shaded darker pink). K-means clustering was therefore applied to determine the within-cluster sums of squares (WCSS) for solutions ranging from 2 to 7 clusters. Visual inspection of the plotted WCSS values (**Figure 2, right panel**) showed a clear inflection point at 5 clusters, indicating that WCSS changed only minimally after 5 clusters. Thus, we considered five clusters to be the optimal solution.

**Figure 2.**
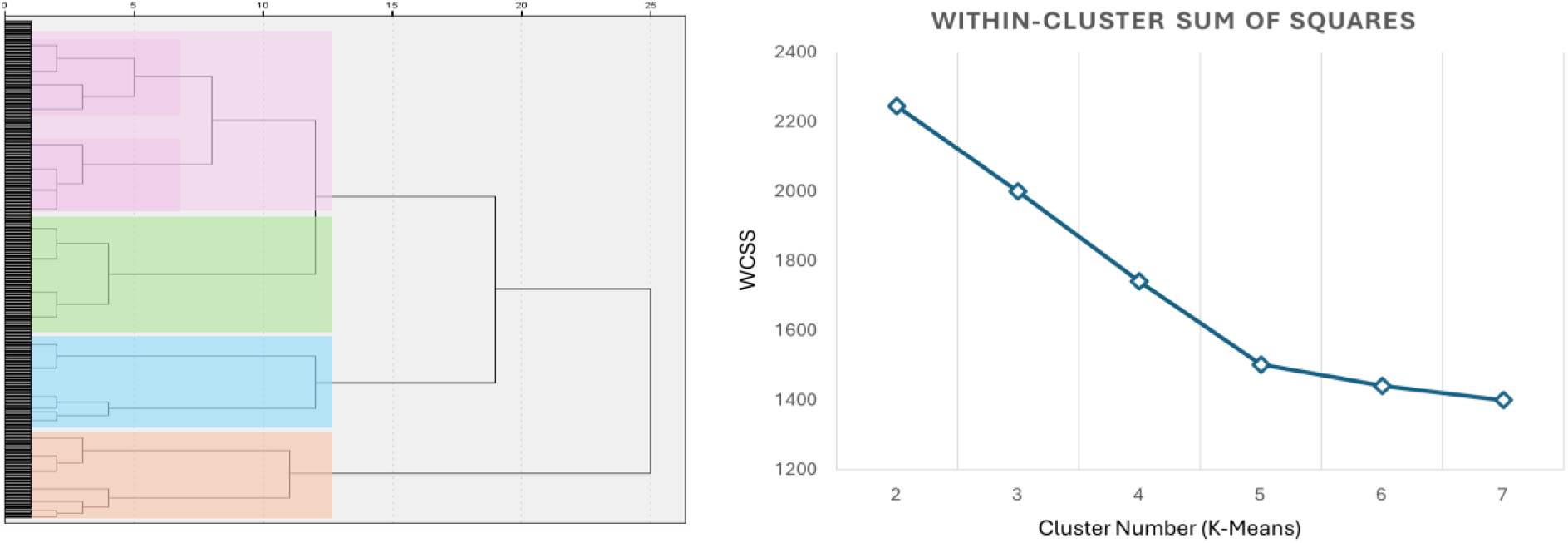
Figure 2 (left panel) shows the dendrogram obtained from the hierarchical agglomerative cluster analysis suggesting either 4 clusters (pink, green, blue and rust) or 5 clusters (including two separate darker shaded clusters within the top pink cluster). The right panel shows the elbow plot for the within-cluster sum of squares (WCSS) for K-Means cluster solutions ranging from 2 to 7 clusters. The inflection point suggests an optimal solution of 5 clusters.

### Cluster solution

Standardized risk category scores were entered into a K-means analysis to determine membership for a 5-cluster solution, using squared Euclidean distances. Convergence was achieved in 10 iterations, with no additional change to the cluster centers from the initial minimum distance between centers. Cluster membership and squared Euclidean distance metrics were saved for each participant. Final cluster centers and N’s for each cluster are provided in **Table 1**, as well as the F statistic for each risk category from the cluster ANOVA, indicating that all five risk categories contributed to the final cluster solution, with F’s(4,550) ranging from 86.19 to 209.47, all p’s <.001.

**Table 1.** Table 1 lists the standardized final cluster centers, expressed as mean Z scores, for each risk category from the K-Means 5-cluster analysis, including the N’s for each resulting cluster and the F(4,550) statistic for each risk category, indicating that all five risk categories contributed significantly to the cluster solution.

|  | Cluster 1 | Cluster 2 | Cluster 3 | Cluster 4 | Cluster 5 |  |
| --- | --- | --- | --- | --- | --- | --- |
|  | N=149 | N=166 | N=75 | N=103 | N=62 |  |
| **Risk Category** |  |  |  |  |  |  |
| Cardiovascular Risk | -1.05 | 0.64 | 0.70 | -0.10 | 0.17 | F=131.09, p<.001 |
| Neuropathology | -0.55 | -0.51 | 0.15 | 1.53 | -0.24 | F=209.47, p<.001 |
| Glucose Dysregulation | -0.34 | -0.37 | 1.76 | -0.28 | 0.17 | F=144.26, p<.001 |
| Inflammation | -0.21 | -0.25 | -0.19 | -0.26 | 1.86 | F=103.71, p<.001 |
| Immune Dysfunction | -0.29 | -0.24 | 0.14 | -0.31 | 1.70 | F=86.19, p<.001 |

### Resulting clusters

The resulting 5 cluster profiles are plotted in **Figure 3** and labelled based on their profile of category risks. To decrease confusion between the risk categories and the cluster labels, clusters will be referred to by cluster number (C1 through C5) along with their descriptive label. The first cluster shown in orange and labelled C1 Low Risk (N=149, 27% of the cohort) had standardized risk scores (expressed as Z) below the mean in all risk categories. The second cluster shown in pink and labelled C2 Cardiovascular (N=166, 30%) only differed from the C1 Low Risk cluster due to their higher mean cardiovascular risk scores, with all other risk scores nearly identical across the two groups and below the mean. The third group, shown in green and labelled C3 Glucose Dysregulation (N=75, 13.5%), showed a prominent peak mean z score for glucose dysregulation with a moderate level of cardiovascular risk. The fourth cluster shown in blue and labelled C4 Neuropathology (N=103, 18.5%) had the highest mean risk score in the neuropathology category relative to all other groups, with mean risks at or below average in all other categories. The final cluster, shown in yellow and labelled C5 Inflammation/Immune Dysfunction (N=62; 11%) had similarly high mean risk scores in both the inflammation and immune dysfunction risk categories. Interestingly, no cluster emerged with a higher mean risk score for cardiovascular risk relative to other groups. Instead, cardiovascular risk was at or above the mean for all risk groups with the exception of the C1 Low Risk cluster, suggesting that cardiovascular risks were spread relatively evenly across four of the five resulting clusters. The heat map (**Figure 4, left panel**) plots the Z scores in the five risk categories (X axis) for every participant, organized by cluster group (Y axis). The map demonstrates a high degree of consistency of the cluster pattern for members within each cluster.

**Figure 3.**
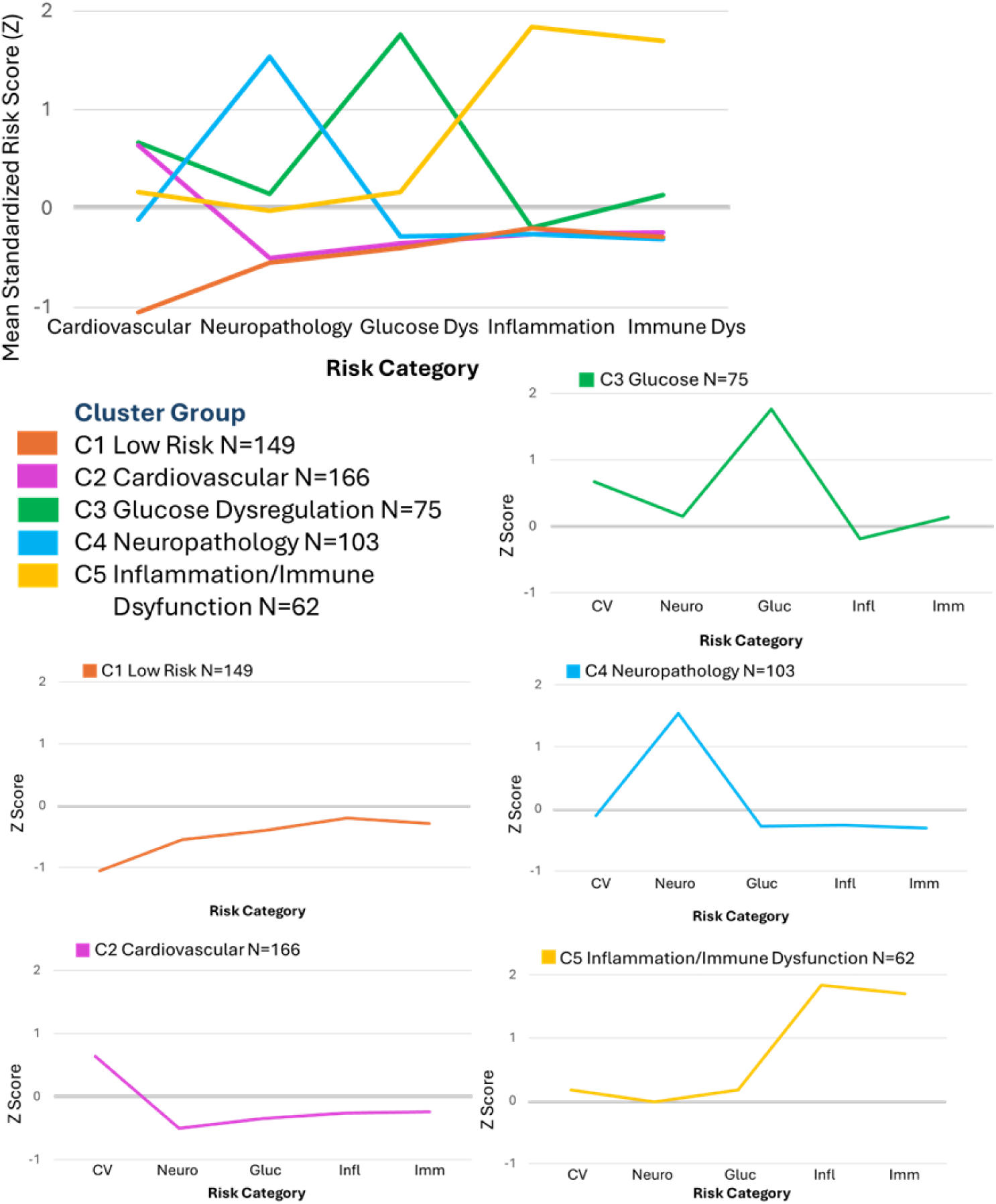
Figure 3 shows the resulting five cluster profiles collectively and separated by cluster, plotting the mean standardized risk category scores (Z scores) within each cluster. Risk categories plotted on the X axis include cardiovascular, neuropathology, glucose dysregulation, inflammation, and immune dysfunction.

**Figure 4.**
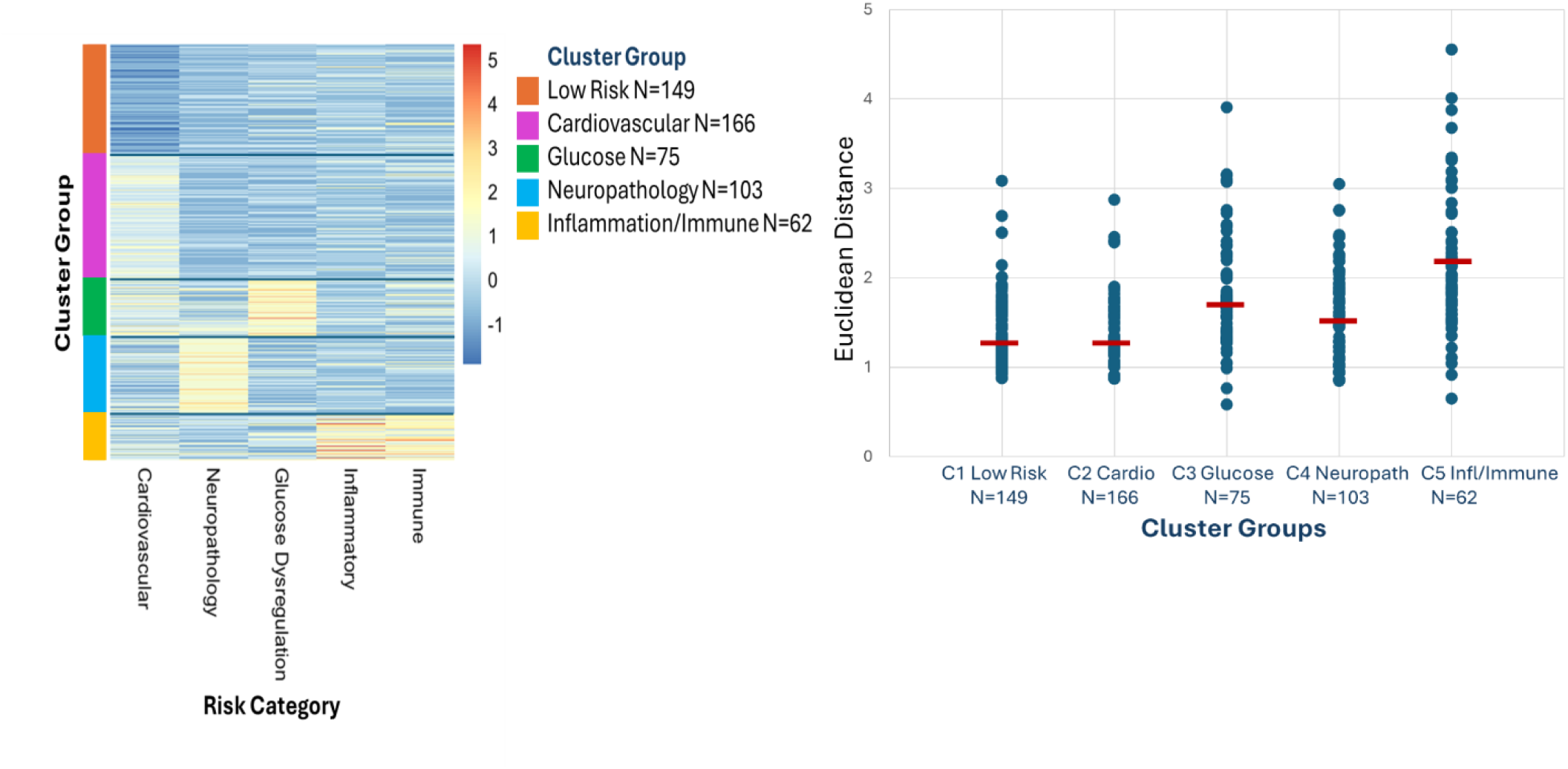
The left panel shows the heat map plotting standardized Z scores for each participant in the five risk categories (X axis), organized by the cluster group along the Y axis. Z scores range from blue (below average risk) to red (above average risk). The map demonstrates a high degree of consistency of the cluster pattern for members within each cluster. In the right panel, Euclidean distances for each member from their respective cluster profile, and the mean Euclidean distance for each cluster indicated by the red bar. Cluster groups include C1 Low Risk, C2 Cardiovascular, C3 Glucose Dysregulation, C4 Neuropathology, and C5 Inflammation/Immune Dysfunction.

### Euclidean Distances

The compactness or cohesion within each cluster is also indicated by the distribution of Euclidean distances from the resulting cluster profile, shown in **Figure 4, right panel**, with lower scores indicating higher similarity between a given individual’s profile and the mean cluster profile. Overall, the clusters show tight grouping of distance measurements for most cluster members, with a handful of outliers in each cluster. One-way ANOVA comparing Euclidean distances across the clusters indicated significant differences across clusters, F(4,550)=48.87, p<.001. In follow-up LSD pairwise t-tests, the C1 Low Risk and C2 Cardiovascular clusters had the lowest mean distances which did not differ significantly from one another, p=.69, indicating that these two clusters profiles were most consistent across members. All other groups differed significantly from the C1 Low Risk and C2 Cardiovascular clusters, all p’s<.001. The C5 Inflammation/Immune Dysfunction cluster had the highest mean distance scores with the most variability relative to all other groups, p’s<.001, likely due in part to the smaller cluster size (62 members). With a larger sample size, it is possible that this cluster would separate further into additional separate groups.

### Contributions of specific factors to cluster groups

To consider which individual factors included in a risk category were most useful in differentiating between clusters, **Table 2** shows the percentage of participants within a cluster with a positive risk score for each factor included in the risk category. The For example, 34% of the C4 Neuropathology group members had a Z score > 1 for ptau 181, while the prevalence for the same factor ranged from 2.4% in the C2 Cardiovascular group to 12.9% in the C5 Inflammation/Immune group. In contrast, the Abeta 42/40 ratio risk was spread more evenly across clusters. Although the prevalence was numerically highest within the C4 Neuropathology cluster (17.5%), it was equally high within the C3 Glucose Dysregulation group (17.3%), with other groups ranging from 6.7% to 9.7%. The pattern suggests that Abeta 42/40 contributed to the clustering, but was not uniquely associated with the C4 Neuropathology cluster.

**Table 2.** Table 2 shows the prevalence for each risk factor within the clusters. The cluster with the highest numerical prevalence rate for each factor is highlighted in blue (with the exception of ‘tobacco use’, where prevalence was nearly identical across four of the five clusters). Bolded percentages indicate risks that were considered to be ‘highly contributing factors’, where the prevalence within one cluster was at least double the prevalence relative to all other clusters.

|  | C1<br>Low Risk<br>N=149 | C2<br>Cardiovascular<br>N=166 | C3<br>Glucose<br>N=75 | C4<br>Neuropathology<br>N=103 | C5<br>Inflammation/<br>Immune<br>N=62 |
| --- | --- | --- | --- | --- | --- |
| **Neuropathology** |  |  |  |  |  |
| ptau 181 | 6.0% | 2.4% | 8.0% | **34.0%** | 12.9% |
| GFAP | 4.7% | 6.6% | 9.3% | **30.1%** | 16.1% |
| NFL | 0.0% | 1.2% | 2.7% | **3.9%** | 3.2% |
| Abeta 42/40 ratio | 6.7% | 7.8% | 17.3% | **17.5%** | 9.7% |
| APOE e4 carrier | 12.8% | 15.1% | 38.7% | **77.7%** | 29.0% |
| PRS Late Onset AD | 2.7% | 3.0% | 22.7% | **65.0%** | 11.3% |
| **Glucose Dysregulation** |  |  |  |  |  |
| Diabetes diagnosis/meds | 2.7% | 5.4% | **58.7%** | 6.8% | 21.0% |
| PRS Type 2 diabetes | 12.8% | 7.2% | **44.0%** | 12.6% | 17.7% |
| PRS Type 1 diabetes | 15.4% | 10.2% | **36.0%** | 11.7% | 16.1% |
| Fasting glucose | 19.5% | 23.5% | **60.0%** | 19.4% | 30.6% |
| Hemoglobin A1C | 0.7% | 1.8% | **41.3%** | 5.8% | 11.3% |
| **Cardiovascular Risks** |  |  |  |  |  |
| Tobacco use | 11.4% | 32.5% | 33.3% | 33.0% | 30.6% |
| Obesity | 12.1% | 42.5% | **52.0%** | 24.3% | 25.8% |
| Hypertension dx/meds | 12.8% | 61.4% | **70.7%** | 41.7% | 62.9% |
| Heart disease diagnosis | 0.0% | 9.6% | 9.3% | 5.8% | 8.1% |
| LDL cholesterol | 9.4% | 21.7% | 17.3% | 12.6% | 8.1% |
| Waist/hip ratio | 39.6% | 87.3% | 84.0% | 64.1% | 79.0% |
| PRS Myocardial infarct | 11.4% | 21.1% | 10.7% | 15.5% | 11.3% |
| PRS hyperlipidemia | 7.4% | 20.5% | **24.0%** | 16.5% | 19.4% |
| CRP | 0.0% | 10.2% | **14.7%** | 2.9% | 8.1% |
| Statin meds | 24.2% | 71.1% | 61.3% | 50.5% | 54.8% |
| **Inflammation** |  |  |  |  |  |
| PRS TNF cytokine | 16.1% | 13.9% | 14.7% | 12.0% | **24.2%** |
| IL 1a | 6.7% | 0.6% | 1.3% | 1.9% | **27.4%** |
| IL 1b | 4.0% | 4.2% | 2.7% | 2.9% | **48.4%** |
| IL 6 | 0.7% | 1.2% | 0.0% | 0.0% | **4.6%** |
| TNFa | 4.7% | 1.8% | 2.7% | 3.9% | **53.2%** |
| TNFb | 2.0% | 3.0% | 1.3% | 1.0% | **32.3%** |
| IL 12 p40 | 1.3% | 0.6% | 0.0% | 1.9% | **40.3%** |
| IL 17a | 4.7% | 2.4% | 6.7% | 9.7% | **32.3%** |
| Anti-inflammatory meds | 16.8% | 23.5% | 29.3% | 17.5% | **32.3%** |
| **Immune risks** |  |  |  |  |  |
| MIP 1a | 4.7% | 1.8% | 8.0% | 1.0% | **38.7%** |
| MIP 1b | 6.0% | 4.8% | 12.0% | 2.9% | **45.7%** |
| IP 10 | 3.4% | 1.8% | 2.7% | 1.0% | **6.5%** |
| MCP 1 | 6.7% | 10.2% | 18.7% | 5.8% | **46.8%** |
| MDC | 8.7% | 11.4% | 16.0% | 10.7% | **35.5%** |
| Eotaxin | 10.7% | 8.4% | 9.3% | 7.8% | **37.1%** |
| Asthma diagnosis/meds | 4.0% | 13.3% | 26.7% | 11.7% | 22.6% |
| PRS Allergy/Asthma/Eczema | 14.8% | 12.0% | 9.3% | 16.3% | 30.6% |

Several patterns were notable from **Table 2**. Within the C3 Glucose dysregulation cluster, all five glucose dysregulation risk factors were high contributors, including diabetes diagnosis/medication use, hemoglobin a1c levels, fasting glucose levels, and PRS for Type 1 and Type 2 diabetes. Interestingly, this group also had the highest prevalence of obesity, hypertension, CRP, and PRS for hyperlipidemia, risk factors that were included in the cardiovascular risk category, although none met the criterion for a highly contributing factor.

Two other clusters were noteworthy. First, the C5 Inflammation/Immune Dysfunction cluster showed strikingly high prevalence rates on virtually all inflammatory and immune measures relative to other groups, as well as high PRS for allergies/asthma/eczema. The only exception was asthma diagnosis/medication use which was equally high in this cluster and the C3 Glucose Dysregulation group. Second, the C1 Low Risk cluster had consistently low prevalence rates in every risk category relative to other clusters, even among the cardiovascular risk factors. While the prevalence of cardiovascular risks tended to be spread relatively evenly across the four high risk clusters, the C1 Low Risk group had prevalence rates that were at least 50% *lower* than all other groups, including the lowest rates of tobacco use, hypertension, heart disease, waist/hip ratios, C-reactive protein values, and use of statin medication. These risk factors also differentiated the C1 Low Risk group from the C2 Cardiovascular group, although the two groups appeared similar to one another in other risk categories.

Polygenic risk scores (PRS) consistently contributed to differentiation between clusters and tracked with other factors within their assigned risk categories. Within the C5 Inflammation/Immune cluster, prevalence rates for PRS’s for cytokine responsiveness and immune conditions (allergies/asthma/eczema) were double the prevalence rates observed in other cluster groups. A similar pattern was observed for PRS’s for Type1 and Type 2 diabetes within the C3 Glucose Dysregulation cluster, the PRS for late onset AD within the C4 Neuropathology cluster, and the PRS for myocardial infarct risk within the C2 Cardiovascular cluster. The only exception was the PRS for hyperlipidemia.

While this PRS was included in the cardiovascular risk category, prevalence rates were similarly high in the C2 Cardiovascular cluster (20.5%) and the C3 Glucose Dysregulation cluster (24%). This may not be surprising given that hyperlipidemia is a risk factor for both cardiovascular diseases and diabetes.

Whether or not the genetic risk scores overlapped with markers of phenotypic expression or other related risk factors depended on the risk cluster. For example, within the C3 Glucose Dysregulation cluster, high PRS for Type 1 and Type 2 diabetes were present in 80% of the cohort (44% and 36%, respectively). However, only 42% reported either a diagnosis of diabetes or the use of diabetes-related medication (such as insulin or metformin). In other cases, PRS were overlapping with other factors within a risk category. For example, 65% of the C4 Neuropathology cluster had a high PRS for late onset AD (n=100). Of these individuals, a high percentage (73%, n=73) were also *APOE* ε4 carriers, likely because the PRS score incorporates *APOE* status. However, it is equally important to note that 27 participants had a high PRS score who were not *APOE* ε4 carriers, again suggesting the PRS may contribute independently to risk beyond other related risk factors or phenotypes.

### Comparing demographics and cognitive scores across the five clusters

Mean age, educational attainment, and sex for the five clusters are listed in **Table 3**. Ages ranged from 50 to 79 in all groups.

**Table 3.** Table 3 shows the N (female, male) for each cluster, the mean and standard deviation (SD) for age (in years) and the mean and standard deviation (SD) for educational attainment for each of the five cluster groups. Educational attainment was coded as follows: 1=Less than high school diploma, 2=High school diploma, 3=Some college, 4=College degree, and 5=Postgraduate degree.

| Cluster Group | N | Age |  | Education |  |
| --- | --- | --- | --- | --- | --- |
|  |  | Mean | SD | Mean | SD |
| C1 Low Risk | 149 [113,36] | 62.22 | 7.06 | 4.15 | 0.86 |
| C2 Cardiovascular | 166 [109, 57] | 66.45<sup>1</sup> | 6.80 | 3.98 | 0.92 |
| C3 Glucose | 75 [48, 27] | 64.45 | 8.04 | 3.92 | 1.12 |
| C4 Neuropathology | 103 [64, 39] | 64.71 | 8.09 | 4.03 | 1.05 |
| C5 Inflammation/Immune | 62 [41,21] | 64.24 | 7.72 | 3.63<sup>2</sup> | 1.11 |

<sup>1</sup> C1 Cardiovascular was older than all other groups, $p's <.05$
<sup>2</sup> C5 Inflammation/Immune had lower educational attainment than all other groups, $p's <.05$

Mean differences in age were below 5 years, ranging from 62.22 (C1 Low Risk) to 66.45 (C2 Cardiovascular). One-way ANOVA for age (in years) indicated a significant difference across groups, F(4,550)=5.06, p<.001. Follow-up t-tests (p’s <.05) indicated that the C2 Cardiovascular group was older than all other groups, with no additional group differences. Educational attainment also differed significantly across the clusters, indicated by a one-way ANOVA, F(4,550)=3.28, p<.05. Follow-up paired t-tests (p’s<.05) indicated that the C5 Inflammatory/Immune cluster had lower educational attainment compared to all other cluster groups, with no other differences among groups. All groups had higher percentages of females compared to males, ranging from 62% to 66% female in the four high risk clusters. Although the percentage of females in the C1 Low Risk cluster was numerically higher (76%), the group differences did not meet statistical significance, χ(4) = 5.64, p=.23, ns.

To control for Type 1 error, the four memory measures from the AVLT were entered into a GLM as a repeated measure, with cluster membership entered as a between-subjects measure, and age and educational attainment entered as covariates. We note that, because the resulting clusters did not differ in the ratio of female to male participants, sex was not included as a covariate in the GLM. As expected, both age and education affected memory performance, F’s(1,548)=41.31 and 20.81, respectively, p’s<.001. Controlling for these variables, memory measures differed across clusters, indicated by a main effect of cluster membership, F(4,548)=2.60, p=.036. Importantly, cluster membership did not interact with memory measure, suggesting that the impact of cluster membership was similar across the memory measures, F(4,548)=1.81, p=.12.

To identify potential differences in memory performance among the cluster groups, we followed the omnibus test with Least Significant Difference pairwise t-tests. Based on the results of the cluster analysis, we hypothesized that the C1 Low Risk group would obtain higher memory scores relative to all other groups with elevated risk scores in one or more risk category. However, we had no clear hypotheses regarding differences between high risk clusters. We therefore report only on pairwise differences between C1 Low Risk and the four high risk clusters. As listed in **Table 4**, the C1 Low Risk group had the highest scores on all four memory measures. For Total Recall and Total Delayed Recall, C1 Low Risk scores were significantly higher than all other groups, t’s > 2.02 and 2.31, respectively, p’s<.05. On Trial 1 Recall, the C1 Low Risk group scores were higher compared to C3 Glucose Dysregulation, C4 Neuropathology, and C5 Inflammation/Immune dysfunction groups, t’s > 1.90, p’s<.05. On Trial 5 Recall, the C1 Low Risk group scores were higher compared to the C3 Glucose Dysregulation and C5 Inflammation/Immune dysfunction groups, t’s > 1.89, p’s<.05.

**Table 4.** Performance for the five cluster groups on four measures from the AVLT, including the number of words recalled on Trial 1 (Trial 1 Recall), words recalled on Trial 5 (Trial 5 Recall), the total words recalled across Trials 1 though 5 (Total Recall), and the total words recalled after a delay (Total Delayed Recall). Listed are the group n’s, the mean and standard error of the mean (SEM), and range (Min, Max) for each cluster group, and p values (two-tailed) for LSD t-tests comparing each group to the C1 Low Risk cluster group, controlling for age and education. *Indicates a significantly lower score relative to C1 Low Risk.

|  | Cluster Group | N | Mean | SEM | Min | Max | p |
| --- | --- | --- | --- | --- | --- | --- | --- |
| Total Recall | **C1 Low Risk** | **149** | **50.58** | **.717** | **27** | **68** |  |
|  | C2 Cardiovascular | 166 | 48.42* | .766 | 20 | 72 | .044 |
|  | C3 Glucose Dysregulation | 75 | 47.65* | 1.188 | 13 | 66 | .029 |
|  | C4 Neuropathology | 103 | 48.11* | .874 | 23 | 69 | .042 |
|  | C5 Inflammatory/Immune | 62 | 45.19* | 1.268 | 26 | 64 | <.001 |
| Trial 1 Recall | **C1 Low Risk** | **149** | **6.03** | **.144** | **2** | **11** |  |
|  | C2 Cardiovascular | 166 | 5.70 | .134 | 1 | 13 | .197, ns |
|  | C3 Glucose Dysregulation | 75 | 5.43* | .219 | 0 | 9 | .015 |
|  | C4 Neuropathology | 103 | 5.61* | .178 | 1 | 11 | .045 |
|  | C5 Inflammatory/Immune | 62 | 5.40* | .180 | 3 | 9 | .018 |
| Trial 5 Recall | **C1 Low Risk** | **149** | **12.50** | **.170** | **6** | **15** |  |
|  | C2 Cardiovascular | 166 | 12.11 | .175 | 5 | 15 | .144, ns |
|  | C3 Glucose Dysregulation | 75 | 12.04 | .300 | 3 | 15 | .168, ns |
|  | C4 Neuropathology | 103 | 11.93* | .226 | 6 | 15 | .05 |
|  | C5 Inflammatory/Immune | 62 | 11.15* | .336 | 5 | 15 | <.001 |
| Total Delayed Recall | **C1 Low Risk** | **149** | **10.42** | **.246** | **0** | **15** |  |
|  | C2 Cardiovascular | 166 | 9.60* | .271 | 0 | 15 | .030 |
|  | C3 Glucose Dysregulation | 75 | 9.45* | .41 | 0 | 15 | .041 |
|  | C4 Neuropathology | 103 | 9.60* | .315 | 1 | 15 | .046 |
|  | C5 Inflammatory/Immune | 62 | 8.73* | .465 | 1 | 15 | <.001 |

## Discussion

In summary, we observed four clusters of older adults with profiles that highlighted one or more areas of risk that are known to negatively impact cognitive aging, and a separate cluster, comprising 27% of the cohort, with below-average risks in all categories relative to the full sample. Importantly, consistent with our hypothesis, the four high risk profiles were associated with poorer performance on multiple memory measures from a widely-used neuropsychological memory test, relative to the low risk group.

The five risk categories included here were chosen specifically because of their known impact on cognitive aging. Unique to the present study was the observation of clear separation between the risk profiles among a large group of older adults without a diagnosis of dementia. While some individuals within the clusters might be classified as ‘mild cognitive impairment’ (MCI) due to their lower memory scores, the causes of MCI are multi-factorial, and may not always be due to neurodegenerative disease. MCI should be considered a risk for dementia, given that most individuals with MCI will not develop dementia in their lifetime (Petersen et al., 2014; Salemme et al., 2025). The current results highlight the importance of taking an individualized approach – a *Precision Aging* approach – for characterizing for age-related cognitive impairment. Lower memory performance relative to the low risk group was evident in multiple groups of older adults, but possibly for different underlying reasons.

Other studies have employed data-driven clustering methods in determining the best predictors of cognitive aging. For example, Chew et al. (2021) employed k-means clustering to identify subgroups with different profiles of functional abilities associated with poorer cognitive performance, more depressive symptoms, and declines in physical functioning over a one-year period. Djonlagic et al. (2021) used hierarchical density-based clustering to identify patterns of sleep disturbances that were associated with poorer cognition and other negative health outcomes such as depression and diabetes. Using k-means clustering, Roca-Ventura et al. (2025) identified complex profiles of physical, psychological, social, and health-related factors that were differentially associated with poorer cognition and cognitive decline.

The present study builds on this approach by considering multiple factors associated with separate domains of risk within the same cohort. We demonstrated the utility of composite scores for predicting risk, rather than considering single factors. As evident from **Table 4**, the total number of risks within each category, rather than any single risk factor, determined cluster membership. Most risk factors contributing to cluster membership had prevalence rates ranging from 20-35%; fewer than 25% of the individual factors had prevalence rates above 50%. Composite scores have been used widely in aging research to capture the aggregate impact of multiple risks, most notably for heart disease (Andersson et al., 2019; Del Giorno et al., 2023) and dementia (Rundek et al., 2020), but also capturing individual differences across cognitive domains, such as memory versus executive functioning (Glisky & Kong, 2008; Glisky et al., 2022). In general, composite scores are more consistent and stable compared to single measures (Song et al., 2013; Kane & Case, 2004; McClure et al., 2024). Our results suggest that the total burden of risks, rather than a high value on any single measure, best characterizes cognitive risk. As an example, 30% of the inflammation/immune dysfunction group had 4 risk factors, 60% of the group had between 5 and 8 risk factors, and the remaining 10% had 9 or more risks within the two categories, but no single risk factor dominated.

The importance of considering total risk burden, rather than single risk factors, is consistent with our previous work (Lewis et al., 2021). The study, with 70,000 participants from the online MindCrowd platform (http://MindCrowd.org; Ryan et al. 2025), considered the impact of self-reported cardiovascular risks including smoking (current or past), hypertension, obesity, diabetes, and heart disease on paired associates memory test. Each of the individual risk factors was associated with poorer memory performance, controlling for multiple demographic and other risk factors. However, the best predictor of memory performance was the total number of risks, regardless of which cardiovascular risk factors were included.

In the present study, cardiovascular insufficiency behaved differently than other risk categories. While glucose dysregulation, biomarkers of neuropathology, and inflammation and immune dysfunction clustered into distinct profiles, in contrast, a moderate amount of cardiovascular risk was observed in all four of the high risk groups. The cluster referred to as ‘C2 Cardiovascular’ was most accurately defined by the *absence* of other categories of risk. It is possible that the optimal model for predicting cognitive performance may be the combination of a relatively independent risk domain, such as glucose dysregulation or inflammation, plus the added risk imposed by cardiovascular insufficiency. The present results suggest that some risk categories, like glucose dysregulation, can occur relatively independent of other risk categories, while others, like cardiovascular risk, co-occur with multiple other domains of risk.

The neuropathology cluster group was surprising because of the specificity of the cluster in the relative absence of other risks. Notably, the cluster observed here was not defined by a single factor, but instead included contributions from Abeta 42/40, ptau 18, GFAP, and genetic markers, all factors that have been linked to poorer cognitive performance in older adults (Pettigrew et al., 2023; Soldan et al., 2025). Moreover, Abeta 42/40 was equally prevalent in the C3 Glucose Dysregulation group. More work is needed to understand how these combined risk factors may interact to impact age-related cognitive functioning, and how they might be modified by resilience factors including genomics and lifetime experiences (Barulli and Stern, 2013; Pettigrew et al., 2023; Soldan et al., 2013) or biomarkers associated with maintenance of synaptic functions (Anderson et al., 2025; Oh et al., 2025).

PRS for late onset AD, Type 1 and Type 2 diabetes, and immune conditions (asthma/allergies/eczema) were prevalent within their related risk categories. The degree to which a high PRS impacts cognitive functioning in the absence of the phenotype remains to be determined, given that a high PRS does not guarantee disease development. For example, PRS for Type 2 diabetes has been estimated to explain about 15% of familial relative risk, with lifestyle factors significantly altering the outcome (Kim et al., 2024). In the present study, 80% of the C3 Glucose Dysregulation cluster had a high PRS for Type 1 or Type 2 diabetes, or both. However, only 32% of those individuals reported a diagnosis of diabetes, and another 12% reported the use of metformin. This pattern suggests that both the PRS and the phenotype may contribute to risk for age-related cognitive impairment, but whether these factors are additive or confer differing amounts of risk remains to be determined.

A key strength of this study is the multi-domain, person-centered approach, integrating biological risk domains to identify interpretable profiles associated with memory performance. However, several issues warrant further consideration. Cluster analysis depends on analytic choices; thus, findings should be interpreted as data-driven profiles rather than fixed subtypes. Replication and validation in independent cohorts is needed. Risk profiles based on composite scores improve interpretability but may obscure the key combinations of variables or weighting of variables that account for the greatest amount of variance, allowing for more precise prediction within each risk category. An important question is whether the risk profiles predict change in cognition over time rather than just cross-sectionally; longitudinal data are needed to determine stability and prognostic relevance. In addition, while the present results highlights the impact of risk on memory performance, risk clusters may show specificity in terms of different domains of cognitive functioning. For example, poorer episodic memory may be most strongly associated with neuropathology while other categories of risk, such as inflammation and immune dysfunction, may have stronger associations with information processing speed or executive functions.

Importantly, the individualized approach described here provides a roadmap for evaluating the mechanisms of brain structure and function mediating age-related cognitive functioning. We have argued elsewhere (Ryan et al., 2019; Palmer et al., 2025) that multiple types of risk may be mediated by common brain mechanisms, such as vascular damage undermining brain perfusion and white matter integrity, while others may be more strongly related to synaptic function and neuronal loss over time. More direct measures of brain structure/function related to risks, or in combination with patterns of risk, may lead to better prediction of cognitive outcomes and risk for cognitive decline and identify targets for prevention and intervention strategies for maintaining brain health.

In conclusion, the present study represents an important shift in approach for characterizing risks for age-related cognitive impairment among relatively healthy older adults without dementia. Rather than taking a one-size-fits-all approach, the study highlights the importance of considering multiple risk factors and multiple categories of risks, in order to capture the complexity and heterogeneity associated with age-related cognitive functioning among older adults. Such an approach – a *precision aging* approach – is critically important for optimizing brain and cognitive health across the adult lifespan.

## Methods

### Study Design and Participant Recruitment

The Healthy Minds for Life (HML) study of the Precision Aging® Network (PAN; https://precisionagingnetwork.org/) was designed to take an individualized approach to understanding factors that impact brain aging and risk for, or resilience against, age-related cognitive impairment. This multi-site study includes four in-person clinical sites: the University of Arizona (Tucson, AZ), the University of Miami (Miami, FL), Emory University (Atlanta, GA), and the Johns Hopkins University (Baltimore, MD). Participants meet the following inclusion criteria at study entry: aged 50-79 years of age; self-identify as White, Black, or Hispanic race/ethnicity; English language proficiency; no reported diagnosis of memory loss or dementia; no reported history of psychotic illness including schizophrenia or bipolar disorder; and no contraindications for blood draw or brain magnetic resonance imaging (MRI). Enrollment for this study began in 2022 and is ongoing.

HML participants are recruited through MindCrowd (https://mindcrowd.org), a large, online national research study designed to assess individual differences across the adult lifespan and identify factors influencing cognitive performance and age- and disease-related cognitive decline (Huentelman et al., 2020; Ryan et al., 2025), and through local advertisements, social media and community events at each HML site. Potential HML participants are then screened by phone to confirm eligibility and willingness to participate in the in-person study procedures. All participants provide written informed consent prior to participation in the HML study procedures. All study procedures were approved by the WIRB-Copernicus Group (WCG) Institutional Review Board (IRB Tracking Number: 20220776) and were performed following the WCG IRB-approved protocol and in accordance with the Declaration of Helsinki.

### Study procedures

Full details of the HML protocol and links to all HML data can be found on the PAN website (https://precisionagingnetwork.org/). Briefly, the baseline assessment includes a short neuropsychological battery that consists of the Montreal Cognitive Assessment (MoCA; Nasreddine et al, 2005), the Rey Auditory Verbal Learning Test (AVLT; Rey, 1958), and the North American Reading Test (Nelson & Willison, 1991), screening for depression (Patient Health Questionnaire-9; Kroenke et al., 2001), blood draw, 3T MRI based on the ADNI3 advanced protocol (Gunter et al., 2017), carotid ultrasound imaging, and biometric assessments including blood pressure, height, weight, and waist and hip circumference, among others. Subsets of participants also complete a series of additional online cognitive tests through the MindCrowd website (http://mindcrowd.org), selected from the experimental cognitive aging literature (Ryan et al., 2025) evaluating associative memory, executive functions, and complex reaction time, as well as surveys covering demographics, health and medical history (e.g., medical diagnoses, medications, surgeries), and various quality of life measures.

### Participants

For the present analyses, 555 participants with a mean age of 65 years (SD=7.6, range 50 to 79 years) were selected from the first two years of the HML study with complete data including blood biomarkers, health and medical information, in-person cognitive testing (MoCA, AVLT, NART), and MRI. MoCA scores averaged 26.30 (SD=2.79, range 17 to 30). Although some individuals had MoCA scores that would be considered within the mild-to-moderate cognitively impaired range, no participant reported difficulties in activities of daily living that would suggest a diagnosis of dementia. The ratio of female to males was approximately 2:1 (n=376 or 68% female, n=179 or 32% male). Educational attainment was coded as follows: 1=Less than high school diploma, 2=High school diploma, 3=Some college, 4=College degree, and 5=Postgraduate degree. The cohort was relatively well educated, with 72% (n=403) of the sample reporting a college degree or postgraduate degree. The remaining cohort members reported ‘some college’ (19%), ‘high school diploma’ (6%) or ‘less than a high school diploma’ (2%).

### Risk factors

Individual risk factors available from the HML protocol were selected based on their known association with the five risk categories – inflammation, immune dysfunction, cardiovascular health, glucose dysregulation, and neuropathology – including self-reported diagnoses and/or medication use, blood-based biomarkers, and genetic risks including *APOE* ε4 status and relevant polygenic risk scores (PRS). Circulating cytokines and chemokines were measured using the MILLIPLEX® MAP Human Cytokine Magnetic Bead Panel (MilliporeSigma, Cat# HYCTA60K) in accordance with the manufacturer’s recommended protocol. Neuropathology biomarkers, including neurofilament light chain (NfL), glial fibrillary acidic protein (GFAP), amyloid-beta (Aβ40 and Aβ42), were assessed from plasma using Single Molecule Array (Simoa) Neurology 4-Plex E, with phosphorylated tau (ptau-181) assays on a HD-X instrument (Quanterix Corporation). All assays were run in duplicate. Cardiovascular, metabolic and lipid panels were obtained through clinical laboratory assays (Sonora Quest Laboratories, Arizona).

Genomic DNA was extracted from PAXgene Blood DNA tubes using the Promega Maxwell RSC blood DNA Kit (Cat. #AS1400) according to the manufacturer’s protocol with site-specific preprocessing steps. Prior to genotyping, DNA concentration was assessed using PicoGreen, and samples were normalized to 20 ng/µL for array processing. Genome-wide genotyping was performed using the Illumina Global Screening Array v4.0 following the Illumina Infinium EX assay workflow. Genotype calling and technical review were performed in GenomeStudio.

Polygenic risk scores were calculated following published best-practice guidelines (Marees et al., 2018). Genome-wide association study (GWAS) summary statistics were obtained from the NHGRI-EBI GWAS Catalog (Cerezo et al. 2025) and filtered to retain only biallelic SNPs with Benjamini-Hochberg corrected p values < 0.05. For each trait-specific PRS, effect alleles were harmonized with the imputed genotype data, and weighted scores were computed as the sum of imputed risk allele dosages multiplied by their corresponding GWAS effect sizes. PRS were generated based on the data for late-onset Alzheimer’s disease (AD) GCST002245 (Lambert et al., 2013), Type 2 diabetes GCST007515 (Mahajan et al., 2018), Type 1 diabetes GCST90014023 (Chiou et al., 2021), myocardial infarction GCST011365 (Hartiala et al., 2021), hyperlipidemia using Brunzell criteria GCST90104003 (Trinder et al., 2021), TNF cytokine-related variation GCST90012044 (Folkersen et al., 2020), and immune-related conditions (asthma/hay fever/eczema) GCST005038 (Ferreira et al., 2017).

### Risk scores and composite scores

Individual risk factors were coded as 1 (risk) or 0 (no risk) and then summed to create a composite risk category score. Composite scores have the advantage of weighing multiple risks with a single summary variable and can be more sensitive and robust than single variables (Song et al., 2013). Categories included between 5 and 9 risk factors with a combination of blood-based biomarkers, self-reported diagnoses, medication use, and genetic risks including *APOE e4* status and PRS relevant to each category. The specific factors included in each category and how factors were coded are listed in **Table 1**. Briefly, a self-reported diagnosis and/or medication typically used to treat a diagnosed condition was coded as a risk. For example, hypertension was coded as a risk when a participant reported a diagnosis of hypertension or the use of one or more anti-hypertensive medications. Where available, clinical cutoffs were applied including BMI, waist-to-hip ratio (with different cutoffs for males and females), LDL cholesterol, and fasting glucose. PRS and blood biomarkers were standardized based on the mean and standard deviation of the total sample (n=555), with Z scores >1 coded as a risk. The criterion of Z > 1 was chosen because we are interested in *increased risk*, rather than more extreme criteria that might be used for diagnostic purposes. In cases where low values constitute higher risk (such as Ab42/40 and some PRS scores), the scale was reversed so that Z > 1 always constituted higher risk. For normally distributed biomarkers, Z >1 will result in approximately 16% of the total sample being designated ‘at risk’. However, for biomarkers with skewed distributions, this percentage decreases, because Z scores standardize to a mean of 0 and standard deviation of 1, but they do not change the shape (skewness) of the distribution. For parsimony, all PRS and biomarkers were coded in the same way using standard Z scores, since we do not yet know the most appropriate way to assess what constitutes high risk for each of these metrics.

**Table 1.** Factors included in the five risk categories and how each factor was coded as 0 (no risk) or 1 (risk). Factors were summed within a category to create a ‘risk category score’.

| **Neuropathology** |  |
| --- | --- |
| Phosphorylated tau 181 (ptau181) | Blood biomarker Z-score > 1 |
| Glial fibrillary acidic protein (GFAP) | Blood biomarker Z-score > 1 |
| Neurofilament light protein (NFL) | Blood biomarker Z-score > 1 |
| Amyloid-beta 42/40 ratio (Abeta 42/40) | Blood biomarker Z-score > 1 |
| APOE genotype | One or two copies of the e4 allele |
| PRS late onset Alzheimer's disease | Z-score > 1 |
| **Glucose Dysregulation** |  |
| Diabetes diagnosis/medication | Self-report Type 1 or Type 2 diabetes, or taking a diabetes-related medication |
| PRS Type 2 diabetes | Z-score > 1 |
| PRS Type 1 diabetes | Z-score > 1 |
| Fasting glucose | Blood biomarker Fasting glucose > 100 |
| Hemoglobin A1C | Blood biomarker Z-score > 1 |
| **Cardiovascular** |  |
| Tobacco use | Self-report smoking currently or ever |
| Body Mass Index (BMI) | BMI > 30 |
| Hypertension | Self-report diagnosis or anti-hypertensive medication use |
| Heart Disease | Self-report diagnosis |
| LDL Cholesterol | Blood biomarker Z-score > 1 |
| High waist to hip ratio | Females > 0.85, Males > 0.90 |
| PRS myocardial infarct | Z-score > 1 |
| PRS hyperlipidemia | Z-score > 1 |
| C-reactive protein (CRP) | Blood biomarker Z-score > 1 |
| High Cholesterol | Self-report diagnosis or statin medication use |
| **Inflammation** |  |
| PRS Tumor Necrosis Factor (TNF) Cytokine | Z-score > 1 |
| Interleukin 1a (IL 1a) | Blood biomarker Z-score > 1 |
| Interleukin 1b (IL 1b) | Blood biomarker Z-score > 1 |
| Interleukin 6 (IL 6) | Blood biomarker Z-score > 1 |
| Tumor necrosis factor alpha (TNF a) | Blood biomarker Z-score > 1 |
| Tumor necrosis factor beta (TNF b) | Blood biomarker Z-score > 1 |
| Interleukin 12p40 (IL 12p40) | Blood biomarker Z-score > 1 |
| Interleukin 17 alpha (IL 17a) | Blood biomarker Z-score > 1 |
| Anti-Inflammatory Medication | Self report long-term anti-inflammatory medication use |
| **Immune dysfunction** |  |
| Macrophage Inflammatory Protein 1 alpha (MIP 1a) | Blood biomarker Z-score > 1 |
| Macrophage Inflammatory Protein 1 beta (MIP 1b) | Blood biomarker Z-score > 1 |
| Interferon-gamma-induced protein 10 (IP 10) | Blood biomarker Z-score > 1 |
| Monocyte chemo-attractant protein (MCP 1) | Blood biomarker Z-score > 1 |
| Macrophage-derived chemokine (MDC) | Blood biomarker Z-score > 1 |
| Eotaxin | Blood biomarker Z-score > 1 |
| Asthma | Self report asthma diagnosis or asthma-related medication |
| PRS Asthma Hay Fever or Eczema | Z-score > 1 |

## Analysis methods

### Cluster analysis

Because the objective was to identify individual-level, multi-domain risk profiles, we explicitly used a person-centered, data-driven framework. Cluster analysis is the most direct approach for capturing co-occurring risk configurations without imposing linear or additive assumptions. While latent profile analysis provides model-based alternatives, clustering offers a more transparent and parsimonious framework. Our approach with composite domain risk scores and a two-step clustering procedure ensured robust and interpretable profiles.

To determine the optimal number of clusters, the five risk category scores were first standardized and entered into an agglomerative hierarchical cluster analysis using Ward’s linkage method and squared Euclidean distances. Visual inspection of the resulting dendrogram was used to choose a reasonable number of clusters. K-means clustering was then applied to obtain the within-cluster sums of squares (WCSS) for a range of cluster solutions above and below the cluster number suggested from the dendrogram. WCSS values were plotted and visually inspected to determine the inflection point on the graph indicating where the decrease in WCSS slows, suggesting a good balance between the number of clusters and the compactness of the clusters. The inflection point was considered the optimal solution.

Second, once the optimal number of clusters was identified, the standardized risk category scores were entered into a K-means cluster analysis at the specified cluster number using squared Euclidean distances to determine each participant’s cluster membership. Iterations were set to 99 to achieve full convergence to zero, indicating no additional change in cluster centers from the minimum distance between initial centers. Cluster membership and distance metrics were saved for each participant.

Final cluster centers, N’s for each cluster, and ANOVA F statistics and p values for each risk category contributing to the cluster solution are reported.

### Comparisons across clusters

Resulting clusters were compared on demographics (age, educational attainment, sex) and four measures from the AVLT, including free recall on Trial 1 (Trial 1 Recall), free recall on Trial 5 (Trial 5 Recall), the total free recall across 5 trials (Total Recall), and free recall after a 30- minute delay (Total Delayed Recall). To control for experiment-wise error, the four tests were entered in a general linear model (GLM) as a repeated measure, with cluster membership as a between-subject factor, and age, educational attainment, and sex as covariates. The omnibus test was followed up with one-way ANOVAs for each memory score and Least Significant Difference t-tests were used to identify pair-wise differences among cluster groups.

## Data Availability

All data used in the current study are available online through the Precision Aging Network website (https://precisionagingnetwork.org/).

## Data Availability

All data produced are available online at

https://precisionagingnetwork.org/

## Acknowledgements

This research was funded by the National Institute on Aging grant U19AG065169 (Precision Aging Network; Barnes, PI).

We acknowledge and thank the members of the Precision Aging® Network for their contribution to this research: B. Aimagamabetova, A. Aldabergenova, C. Anderson, M. Albert, C. Babbitt, C. A. Barnes, S. Beres, N. Bhadra, A. Bilgin, Y.F. Bolla, A. Bonfitto, A. Box, R. D. Brinton, E. Burrows, D. Cabral, V. D. Calhoun, S. Callahan, C. Camargo, C. Carrasco, D. Chambers, NK. Chen, Z. Chen, D. Coon, M. M. Crespo, M.D. De Both, W. Degnan III, M. Dehghan Rouzi, K. A. Delgado, A. Dolby, J. Don, V. M. Dotson, K. P. Doyle, K. Ellingson, M. Fan, A. Feal Rodriguez, S. Fox-Rosellini, J.B. Frye, L. F. Gladulich, A. Glinka, L. Gossa, S. Han, M. Hay, S. Hoscheidt, M.J. Huentelman, T. James, K. Johnson, M. Johnson, D. Kartchner, S.Y. Kim, B.J. LaFleur, J.J. Lah, A.J.B. Lee, M. Lee, G. Leito, A.I. Levey, B. Levin, S. Matijevic, M.R. Mehl, N. Merchant, S. Merritt, D. Metz, C.S. Mitchell, M. Modjeski, A. Moghekar, C. H. Na, B. Najafi, M. Naymik, K. Norton, T. Nuno, B. Nursal, E. Paitel, P. Pattany, J. Pekar, C.A. Pettigrew, G. Pfaff, V. Pfeifer, S.E. Roh, T. Rundek, J. R. Runyon, L. Ryan, D. Sama-Borbon, K. Sanders, N. Schork, S. Scott, S. Sharma, A. Sidhu, J. Simon, J. Sloan, T. Smith, S. Sockanathan, A. Sokan, A. Soldan, S. Soto, B. Stark, E. M. Sternberg, X. Sun, M. Swartzlander, F. Taguinod, R. Tandon, M. Taylor John, T. Trouard, C. Ugonna, J.G. Varelo Saboria, H. Venkatachalam, L. White, P. F. Worley, J. Xie, Y. Yang, C. Ye, T. Yuhas, T.K. Zepeda, J. Zhou.

